# A PTX3/LDH/CRP signature correlates with lung injury CTs scan severity and disease progression in paucisymptomatic COVID-19

**DOI:** 10.1101/2021.09.29.21264061

**Authors:** Marco Folci, Enrico Brunetta, Ezio Lanza, Barbara Bottazzi, Alessandro Protti, Gaia Messana, Costanza Lisi, Roberto Leone, Marina Sironi, Elena Generali, Stefano Rodolfi, Michele Sagasta, Antonio Voza, Michele Ciccarelli, Cecilia Garlanda, Luca Balzarini, Alberto Mantovani, Maurizio Cecconi, Humanitas COVID-19 Task Force

**Author notes:** These authors contributed equally: Marco Folci, Enrico Brunetta, Ezio Lanza.

## Abstract

**Background:** Quantitative CT (QCT) analysis is an invaluable diagnostic tool to assess lung injury and predict prognosis of patients affected by COVID-19 pneumonia. PTX3 was recently described as one of the most reliable serological predictors of clinical deterioration and short-term mortality. The present study was designed to evaluate a correlation between serological biomarkers of inflammation and lung injury measured by QCT.

**Methods:** This retrospective monocentric study analysed a cohort of patients diagnosed with COVID-19 and admitted because of respiratory failure, or significant radiological involvement on chest CT scan. All patients, males and non-pregnant females older than 18 years, underwent chest CT scan and laboratory testing at admission. Exclusion criteria were defined by concurrent acute pathological processes and ongoing specific treatments which could interfere with immune activity. The cohort was stratified based on severity in mild and severe forms. Compromised lung at QCT was then correlated to serological biomarkers representative of SARS-CoV-2. We further developed a multivariable logistic model to predict CT data and clinical deterioration based on a specific molecular signature. Internal cross-validation led to evaluate discrimination, calibration, and clinical utility of the tool that was provided by a score to simplify its application.

**Findings:** 592 patients were recruited between March 19th and December 1st, 2020. Applying exclusion criteria which consider confounders, the cohort resulted in 366 individuals characterized by 177 mild and 189 severe forms. In our predictive model, blood levels of PTX3, CRP and LDH were found to correlate with QCT values in mild COVID-19 disease. A signature of these three biomarkers had a high predictive accuracy in detecting compromised lungs as assessed by QCT. The score was elaborated and resulted representative of lung CT damage leading to clinical deterioration and oxygen need in mild disease.

**Interpretation:** The LDH, PTX3, CRP blood signature can serve as a strong correlate of compromised lung in COVID-19, possibly integrating cellular damage, systemic inflammation, myeloid and endothelial cell activation.

**Funding:** This work was supported by a philanthropic donation by Dolce & Gabbana fashion house (to A.M., C.G.) and by a grant from Italian Ministry of Health for COVID-19 (to A.M. and C.G.).

**Research in context:** *Evidence before this study:* Besides nasopharyngeal swab and serological test, chest CT scan represents one of the most useful tools to confirm COVID-19 diagnosis; moreover, QCT has been demonstrated to foresee oxygen need as well as deterioration of health status. Several clinical and serological parameters have been studied alone or combined in scores to be applied as prognostic tools of SARS-CoV-2 pneumonia; however, no one has yet reached the everyday practice. Recently, our group has investigated the expression and clinical significance of PTX3 in COVID-19 demonstrating the correlation with short-term mortality independently of confounders. The result was confirmed by other studies in different settings increasing evidence of PTX3 as a strong biomarker of severity; noteworthy, a recent report analysed proteomic data with a machine learning approach identifying age with PTX3 or SARS-CoV-2 RNAemia as the best binary signatures associated to 28-days mortality.

*Added value of this study:* The present study was designed to investigate associations between markers of damage and the CT extension of SARS-CoV-2 pneumonia in order to provide a biological footprint of radiological results in paucisymptomatic patients. QCT data were considered in a binary form identifying a threshold relevant for clinical deterioration, as already proved by literature. Our findings demonstrate a significant correlation with three peripheral blood proteins (PTX3, LDH and CRP) which result representative of COVID-19 severity. The study presents a predictive model of radiological lung involvement which performs with a high level of accuracy (cvAUC of 0·794±0·107; CI 95%: 0·74–0·87) and a simple score was provided to simplify the interpretation of the three biomarkers. Besides additional finding on PTX3 role in SARS-CoV2 pathology, its prognostic value was confirmed by data on clinical deterioration; indeed, paucisymptomatic subjects showed a 11·9% deaths. The model offers the possibility to quickly assess patients resulted positive for SARS-CoV-2 and estimate people at risk of deterioration despite normal clinical and blood gases analysis, with potential to identify those who need better clinical monitoring and interventions.

*Implications of all the available evidence:* Predicting the extension, severity, and clinical deterioration in COVID-19 patients its pivotal to allocate enough resources in emergency and to avoid health system burden. Despite the urgent clinical need of biomarkers, SARS-CoV-2 pneumonia still lacks something able to provide an easy measure of its severity. Some multiparametric scores have been proposed for severe COVID-19 and rely on deep assessment of patients status (clinical, serological, and radiological data). Our model represents an unprecedented effort to provide a tool which could predict CT pneumonia extension, oxygen requirement and clinical deterioration in mild COVID-19. Based on the measurement of three proteins on peripheral blood, this score could improve early assessment of asymptomatic patients tested positive by SARS-CoV2 specifically in first level hospitals as well in developing countries.

## Introduction

The new SARS-CoV-2 led to the current pandemic of COVID-19, a respiratory disease that ranges from mild conditions^1^ to severe cases, both characterized by different grades of interstitial pneumonia.^2,3^ The most life-threatening manifestations refer to acute respiratory distress syndrome (ARDS) and systemic complications, which can worsen till a multiorgan failure state.^4–7^ By interacting with ACE2 expressed by pneumocytes and endothelial cells, SARS-CoV-2 induces alveolar disruption, endothelial inflammation, and a procoagulant state which ultimately results in respiratory distress.^5,8,9^ The respiratory tract serves as a port-of-entry of SARS-CoV-2 and pneumonia is a major step in disease progression. Chest computed tomography (CT) is a highly sensitive tool used for diagnosis and monitoring of COVID-19.^10–12^ Widely adopted in respiratory affections, CT findings vary depending on the onset of disease: ground-glass opacities and parenchymal consolidations seem to be the most representative of SARS-CoV-2 infection.^13,14^ Even if chest CT is not able to distinguish between viral pneumonia,^15^ the extension of the pathological lung lesions has been associated with its clinical course.^16,17^ Moreover, computer-aided quantitative analysis of the CT exam (QCT) has been related to oxygens needs, orotracheal intubation and unfavourable outcome.^17^ Circulating biomarkers of inflammation serve as invaluable tools to assess disease severity and progression. These include acute phase proteins (e.g. C reactive protein, CRP), cytokines (IL-6), products of the coagulation cascade (fibrin dimer) and cellular enzymes (e.g. lactate dehydrogenase, LDH). Recently we found that high levels of pentraxin-3 (PTX3) measured early upon hospitalization were a strong, independent predictor of unfavourable outcome (death).^18^ Similar results were obtained by others.^18–21^ PTX3 is a distant relative of CRP and a key component of innate immunity, inflammation, and tissue repair.^22–24^ Released at the site of inflammation by epithelia and immune cells under the stimulus of proinflammatory cytokines, PTX3 acts as a soluble pattern recognition molecule conferring local resistance to specific pathogens.^22^ It has been demonstrated a rapid rise in PTX3 plasma levels with fungal, bacterial, and viral infections;^25–27^ indeed, systemic concentration has been associated with severity and mortality in sepsis.^28,29^ Furthermore, PTX3 production by endothelial cells under inflammatory conditions have shed in light the role of this molecule as a marker of disease activity in atherosclerosis and vasculitis.^30–33^ In COVID-19, myelomonocytic cells and lung endothelial cells were major sources of PTX3.^18^ The strong prognostic significance of PTX3 may reflect integration of myeloid cells and lung endothelial cell activation.^31,33^ The present study was designed to assess how blood levels of PTX3, and other biomarkers reflect lung involvement in COVID-19 as measured by QCT. In a retrospective series of 177 non-severe COVID-19 patients, PTX3 results associated with parenchyma injury in univariate and multivariate analysis. A composite score which included LDH, CRP and PTX3 was related with high accuracy to pulmonary alterations at QCT. The strong predictive significance of these biomarkers even in paucisymptomatic patients may reflect integration of tissue damage (LDH), systemic response to inflammation (CRP) and local myelomonocytic and endothelial cell activation (PTX3).

## Material and methods

### Study design and participants

This retrospective study analyzed a cohort of 592 patients. We included all males and non-pregnant females, 18 years of age or older, admitted to Humanitas Clinical and Research Center (Rozzano, Milan, Italy) between March 19th and December 1st, 2020 with a laboratory-confirmed diagnosis of COVID-19. Hospital admission criteria were based on a positive assay for SARS-CoV-2 associated with respiratory failure requiring oxygen therapy, or radiological evidence of significant pulmonary infiltrates on chest computed tomography (CT) scan, or reduction in respiratory/cardiopulmonary reserve as assessed by 6 minutes walking test, or due to frailty related with patient comorbidity. All patients underwent laboratory testing at hospital admission and chest CT scan in the first two days. The outcome of the study was to demonstrate correlations between compromised lung volume at CT scan in COVID-19 patients and serological biomarkers representative of the infection, in particular with PTX3 plasma levels at the admission. Once identified confounders for the study, patients were excluded from the analysis because of concomitant bacterial infections (79, 13·3%), active neoplasia (54, 9·1%), acute myocardial infarction, and stroke. Since the continuously evolving situation in pharmacological management of COVID-19, subjects treated with steroids, low molecular weight heparin (LMWH), and immunosuppressant (total of 54, 9·1%) before the hospital admission were not included. Patients undergone to CT scan after 2 days were excluded (39, 6·6%) since radiological features were not considered representative of the clinical and blood tests assessment. Based on these criteria, the cohort is characterized by 366 individuals (226 excluded), 137 (37·5%) females and 229 (62·5%) males, with a mean age of 69±14·8 years and a median number of days from symptoms onset to the ER admission of 6·2±5·3 days (Supplementary Figure S1). Comorbidities were assessed at admission and presented in Table 1A, instead severity-related biomarkers, laboratory characteristics and inflammation molecules are presented in supplementary (Supplementary Table S1). As of April 30th 2021, all the patients had been discharged or died, while the composite endpoint of clinical deterioration occurred in 114 patients (30·8%), including 59 (16·12%) who were admitted to the ICU, and 63 (17·21%) who died. Of the 59 patients admitted to the ICU, all had been discharged, and 8 (12·7%) had died.

**Table 1A.** Cohort baseline characteristic. Demographics and comorbidities are presented as follows: 1) First column: total cohort of patients (366 subjects); 2) Second column: MILD COVID-19, characterized by paucisymptomatic patients with PaO2/FiO2 ratio greater than 300 (177 subjects) sub-clustered based on QCT compromised lung threshold of 9·5%; 3) Third column: SEVERE COVID-19, symptomatic patients with PaO2/FiO2 ratio fewer than 300 (189 subjects) all the subjects present QTC compromised lung over the selected threshold.

| Variables | Total Cohort<br>(n = 366) | MILD COVID-19<br>PaO2/FiO2≥300 (n = 177) |  |  | SEVERE COVID-19<br>PaO2/FiO2<300 (n = 189) |
| --- | --- | --- | --- | --- | --- |
|  |  | Overall | QTC compromised lung < 9.5% (n = 91) | QTC compromised lung ≥ 9.5% (n = 86) | QTC compromised lung ≥ 9.5% (n = 189) |
| Demographic Characteristics |  |  |  |  |  |
| Mean ± SD; Median (IQR) or n (%) |  |  |  |  |  |
| Age (years) | 69±14.8; 72 (58–81) | 67.8±15.5; 70 (57–81) | 66.6±15.1; 70 (55–78) | 68.96±15.9; 69 (58–82) | 70.09±14.0; 72 (59–81) |
| Gender |  |  |  |  |  |
| Female | 137 (37.5) | 67 (38) | 33 (36) | 34 (39.5) | 70 (37) |
| Male | 229 (62.5) | 110 (62) | 58 (64) | 52 (60.5) | 119 (63) |
| Comorbidities |  |  |  |  |  |
| Hypertension | 198 (54) | 93 (52.5) | 51 (56) | 42 (49) | 105 (55.6) |
| CKD | 33 (9) | 20 (11.3) | 10 (11) | 10 (12) | 13 (6.9) |
| COPD | 29 (8) | 12 (7) | 6 (7) | 6 (7) | 17 (9) |
| Asthma | 18 (5) | 9 (5) | 6 (7) | 3 (3.5) | 9 (4.8) |
| Diabetes | 77 (21) | 40 (22.6) | 23 (25) | 17 (19.8) | 37 (19.6) |
| Dementia | 27 (7.4) | 15 (8.5) | 5 (5.5) | 10 (12) | 12 (6.3) |
| Heart Failure | 29 (8) | 11 (6) | 7 (8) | 4 (4.7) | 18 (9.5) |
| Coronary Artery Disease | 62 (17) | 27 (15) | 14 (15.4) | 13 (15) | 35 (18.5) |
| Cirrhosis * | 7 (2) | 5 (3) | 0 (0) | 5 (5.8) | 2 (1.1) |
| History of neoplasia * | 9 (2.5) | 5 (3) | 5 (5.5) | 0 (0) | 4 (2) |

\* Statistical significance between MILD COVID-19 subgroups (QCT< and ≥ 9.5%): \* p<0.05; \*\* p<0.01; \*\*\* p<0.001.

The study was evaluated by the Ethics Committee of the IRCCS Istituto Clinico Humanitas (independent organization with registered office in Rozzano, Milan-Italy).

The Independent Ethics Committee approved the study (authorization 233/20) and the requirement for informed consent was waived.

### Laboratory test, demographic, and medical history

Laboratory testing at hospital admission included: complete blood count, renal and liver function (transaminase, total/direct/indirect bilirubin, gamma-glutamyl transferase, alkaline phosphatase), creatinine kinase, lactate dehydrogenase (LDH), myocardial enzymes, electrolytes, and triglycerides. A panel of acute phase reactants including interleukin-6 (IL-6), serum ferritin, D-dimer, C-reactive protein (CRP), fibrinogen, and procalcitonin (PCT) was performed. Body temperature, blood pressure, heart rate, peripheral saturation, and respiratory rate were measured in all patients. Arterial blood gas analysis was performed in the emergency department and PaO2/FiO2 was calculated. In all patients, all blood tests and PTX3 were measured within the first few days after the admission date (mean 0·05±0·2 days). Pneumococcal and Legionella urinary antigen tests were routinely performed. Nasopharyngeal swab for influenza A, B and H1N1 were also routinely performed to exclude co-infections. Additional microbiological tests were performed to exclude other pathogens as possible etiological agents when suggested by clinical conditions (bacterial cultures of sputum, blood, and urine). We obtained a comprehensive present and past medical history from patients focusing on those main categories such as hypertension, chronic kidney disease (CKD), chronic obstructive pulmonary disease (COPD), asthma, diabetes, dementia, heart failure (HF), coronary artery disease, cirrhosis, and history of neoplasia. Positivity was assessed based on reverse-transcriptase–polymerase-chain-reaction (RT-PCR) assay for SARS-CoV-2 on a respiratory tract sample tested by our laboratory, in accordance with the protocol established by the WHO (https://www.who.int/emergencies/diseases/novel-coronavirus-2019/technical-guidance/laboratory-guidance). Due to the high false-negative rate of RT-PCR from the pharyngeal swab, two different swabs were performed in every patient to increase the detection rate.^34^ In cases of a negative assay in throat-swab specimens, but with suggestive clinical manifestations, presence of contact history or suggestive radiological evidence for COVID-19, the detection was performed on bronchoalveolar lavage fluid (BAL) or endotracheal aspirate, which has higher diagnostic accuracy. All demographics, medical history and laboratory tests were extracted from electronic medical records and were checked by a team of three expert physicians.

### Chest CT scan and Quantitative analysis

All patients received a noncontrast chest CT scan upon hospital admission, according to an internally approved diagnostic pathway for the pandemic management. All scans were performed in the first days from admission with a mean of 0·17 ± 0·51 days. CT images were processed using a semi-automated software (3D-Slicer with Chest Imaging Platform).^17,35^ Lung volumes were outlined by 3D-Slicer and manually perfected by a radiologist with ten years’ experience in chest CT (E.L.) and two radiology residents (G.M; C.L); then were divided according to the different density into four subvolumes: non-aerated lung (%NNL, density between 100, − 100 Hounsfield Units, HU), poorly aerated (%PAL, − 101, − 500 HU), normally aerated (%NAL, − 501, − 900 HU), and hyperinflated (− 901, − 1000 HU).^36,37^ The additional volume “compromised lung” (%CL) was considered the sum of %PAL and %NNL (− 500, 100 HU) and considered in a binary form based on the 9·5% threshold which is representative of clinical deterioration and oxygen need, as previously demonstrated.^17^

### Sample collection and PTX3 measurement

Venous blood samples were collected during the first five days after hospital admission (mean 0·073±0·26 days), centrifuged, and EDTA plasma was stored at -80°C until use. PTX3 plasma levels were measured, as previously described, by a sandwich ELISA (detection limit 0·1 ng/mL, inter-assay variability from 8 to 10%) developed in-house, by personnel blind to patients’ characteristics.^18^ In each analytical session a sample obtained from a pool of plasma from healthy donors was used as internal control. The mean PTX3 concentration measured in this sample was 1·81+0·9 ng/mL (mean ± SD).

### Statistical methods

Demographic, clinical, laboratory and outcome data were obtained from electronic medical records and patient chart notes using a standardized data collection form. Descriptive statistics included means with standard deviations (SD) and medians with interquartile ranges (IQR) for continuous variables, and frequency analyses (percentages) for categorical variables. Because of non-linearity, PTX3, CRP and IL-6 were used in the logarithmic scale for regression analysis.

Radiological lung injury was considered in a binary form identifying a threshold of 9·5% which was demonstrated to be significant for clinical deterioration and oxygen need.^17^

The cohort of patients was divided into subgroups representative of clinical deterioration and based on PaO2/FiO2 levels measured at admission. The cut-offs refer to the Berlin classification of ARDS^38^ and lead to identify paucisymptomatic patients as those with a PaO2/FiO2 ratio greater than 300 (MILD COVID-19). All the three subgroups with a PaO2/FiO2 ratio under 300 (SEVERE COVID-19) have not been included in the analysis since in these patients the radiological damage was over the chosen threshold of 9·5% preventing the application of the selected statistic.

### Model development

Univariable and multivariable association between “risk factors” and radiological lung injury (presence of QCT damage ≥9·5%) were estimated by means of logistic regression and the magnitude of the association expressed by means of Odds Ratios (OR).

The analyses were based on non-missing data (missing data not imputed). We followed a standard approach for model selection. In the univariable logistic analysis, a criterion of p≤ 0·10 was used to identify candidate predictors. Additionally, variables were selected according to a review of the literature and consensus opinion by an expert group of physician and methodologists. Then, we fitted the multivariable model and used a backwards selection procedure to eliminate those variables not significant in the multivariable framework. The criterion of p ≤ 0·05 was used for determining which one to eliminate. The OR were presented with their 95% confidence interval (CI) and respective *p*-value.

### Model validation

In order to evaluate the goodness of fit of the model and to generate a more realistic estimate of predictive performance a K-fold cross-validation was implemented (cvAUC). Area under the curve (AUC) was estimated iteratively for k samples (the “test” samples) that were independent of the sample used to predict the dependent variable (the “training” sample).

### Score

Finally, we developed a preliminary prognostic index for radiological lung injury in COVID-19 by converting the beta coefficients from the multivariable model to integer values, while preserving monotonicity and simplicity. We compute the cvAUC with its 95% CI to measure the predictive power of our prognostic index. This analysis was also performed in our initial population including septic and neoplastic patients previously excluded (226pts).

Stata 15.0 software was used to analyze the data (Stata Corp., College Station, TX, USA). P-values less than 0·05 were considered statistically significant. All tests were two-sided. No adjustment for multiplicity was applied.

## Results

### Population and PTX3 results

The study analyzed 177 subjects affected by a mild form of COVID-19 (WHO classification, PaO2/FiO2≥300) and admitted to Humanitas Research Hospital between March 19th and December 1st 2020. The cohort is characterized by 67 (38%) females and 110 (62%) males, with a mean age of 67·8±15·5 (Table 1A). Comorbidities were assessed identifying 93 (52·5%) patients affected by hypertension, 20 (11·3%) with chronic kidney disease (CKD), 12 (7%) COPD patients, 9 (5%) with asthma, 40 (22·6%) diabetics and 11 (6%) classified as HF (Table 1A). The median number of days from symptoms onset to the ER admission were 5 (IQR 2-8). The patients analyzed presented the well-described COVID-19 alteration at blood tests. We observed a reduction of peripheral lymphocyte count (1·01±0·5×10 □/L; normal range 1·0–4·0) as well as an undetectable eosinophils level, an increase in LDH (310·6±115·5 U/L; normal range < 248), Ferritin (537·3±585·9 ng/mL; normal range 23·9–336·2), and CRP (7·9±7·1 mg/dL; normal range < 1·0). PTX3 blood levels were significantly elevated in all COVID-19 patients compared to healthy subjects, as already shown in our previous study.^18^ The mean PTX3 concentration in the present cohort was 37·4±44·96 ng/mL at the admission. Alterations in D-dimer (176 subjects) and fibrinogen were assessed. PCT was measured in order to rule out bacterial superinfections.

All clinical and laboratory data were stratified by QCT injury which was considered in a binary form with a threshold of 9·5% (Table 1B). The two groups were analyzed identifying O2 saturation (p=0·003) and PaO2/FiO2 ratio (p=0·0496) as the only relevant difference among severity-related biomarkers, even though the same clinical presentation. Focusing on blood parameters, patients with a worse QCT presented a severer COVID-19 inflammatory profile as demonstrated by enhanced lymphocytopenia (p=0·04) associated to higher levels of white blood cells (p=0·0310), Neutrophils (p=0·0097), LDH (p<0·0001), Fibrinogen (p=0·0038), CRP (p<0·0001) and PTX3 (p<0·0008). Notably, age, gender, comorbidities, days of onset and IL-6 levels did not vary (Table 1B).

**Table 1B.**
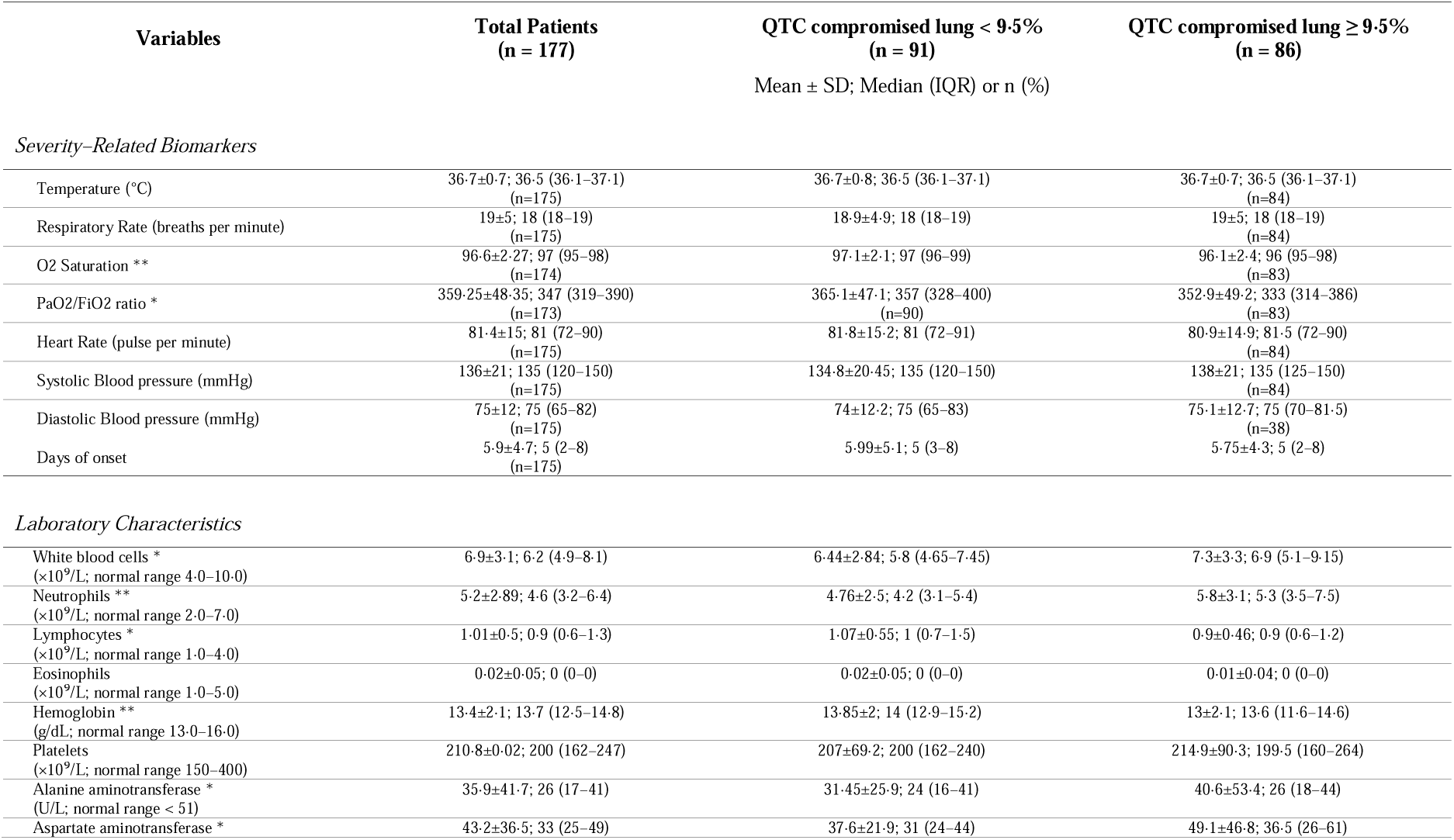
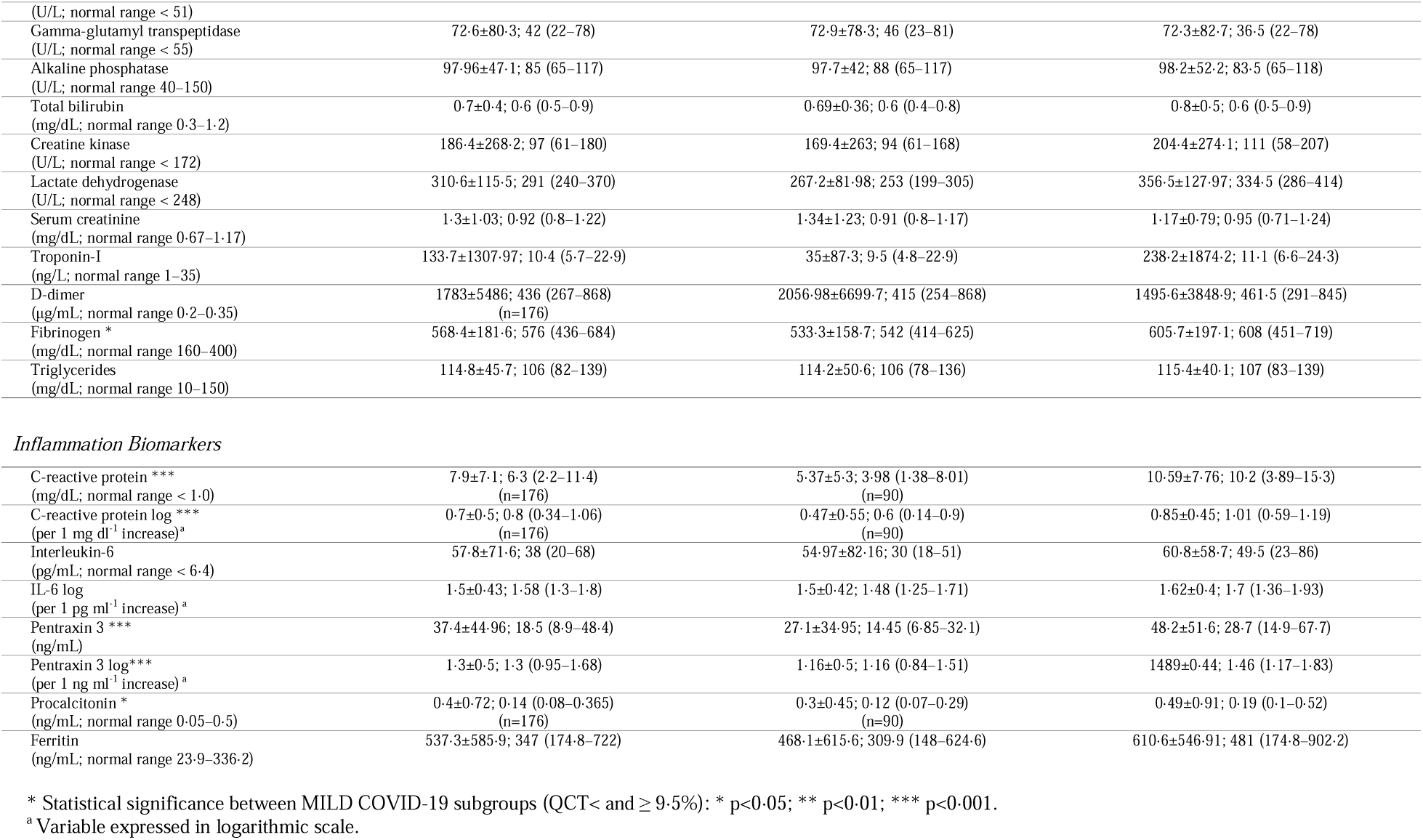
MILD COVID-19 Clinical features and blood tests at admission. Severity-related biomarkers and blood tests are presented for MILD COVID-19 (paucisymptomatic patients with PaO2/FiO2 ratio greater than 300). The two other columns refer to subgroups clustered based on QCT compromission threshold of 9·5%.

| Variables | Total Patients<br>(n = 177) | QTC compromised lung < 9.5%<br>(n = 91) | QTC compromised lung ≥ 9.5%<br>(n = 86) |
| --- | --- | --- | --- |
|  |  | Mean ± SD; Median (IQR) or n (%) |  |
| Severity-Related Biomarkers |  |  |  |
| Temperature (°C) | 36.7±0.7; 36.5 (36.1–37.1)<br>(n=175) | 36.7±0.8; 36.5 (36.1–37.1) | 36.7±0.7; 36.5 (36.1–37.1)<br>(n=84) |
| Respiratory Rate (breaths per minute) | 19±5; 18 (18–19)<br>(n=175) | 18.9±4.9; 18 (18–19) | 19±5; 18 (18–19)<br>(n=84) |
| O2 Saturation ** | 96.6±2.27; 97 (95–98)<br>(n=174) | 97.1±2.1; 97 (96–99) | 96.1±2.4; 96 (95–98)<br>(n=83) |
| PaO2/FiO2 ratio * | 359.25±48.35; 347 (319–390)<br>(n=173) | 365.1±47.1; 357 (328–400)<br>(n=90) | 352.9±49.2; 333 (314–386)<br>(n=83) |
| Heart Rate (pulse per minute) | 81.4±15; 81 (72–90)<br>(n=175) | 81.8±15.2; 81 (72–91) | 80.9±14.9; 81.5 (72–90)<br>(n=84) |
| Systolic Blood pressure (mmHg) | 136±21; 135 (120–150)<br>(n=175) | 134.8±20.45; 135 (120–150) | 138±21; 135 (125–150)<br>(n=84) |
| Diastolic Blood pressure (mmHg) | 75±12; 75 (65–82)<br>(n=175) | 74±12.2; 75 (65–83) | 75.1±12.7; 75 (70–81.5)<br>(n=38) |
| Days of onset | 5.9±4.7; 5 (2–8)<br>(n=175) | 5.99±5.1; 5 (3–8) | 5.75±4.3; 5 (2–8) |
| Laboratory Characteristics |  |  |  |
| White blood cells *<br>(×10<sup>9</sup>/L; normal range 4.0–10.0) | 6.9±3.1; 6.2 (4.9–8.1) | 6.44±2.84; 5.8 (4.65–7.45) | 7.3±3.3; 6.9 (5.1–9.15) |
| Neutrophils **<br>(×10<sup>9</sup>/L; normal range 2.0–7.0) | 5.2±2.89; 4.6 (3.2–6.4) | 4.76±2.5; 4.2 (3.1–5.4) | 5.8±3.1; 5.3 (3.5–7.5) |
| Lymphocytes *<br>(×10<sup>9</sup>/L; normal range 1.0–4.0) | 1.01±0.5; 0.9 (0.6–1.3) | 1.07±0.55; 1 (0.7–1.5) | 0.9±0.46; 0.9 (0.6–1.2) |
| Eosinophils<br>(×10<sup>9</sup>/L; normal range 1.0–5.0) | 0.02±0.05; 0 (0–0) | 0.02±0.05; 0 (0–0) | 0.01±0.04; 0 (0–0) |
| Hemoglobin **<br>(g/dL; normal range 13.0–16.0) | 13.4±2.1; 13.7 (12.5–14.8) | 13.85±2; 14 (12.9–15.2) | 13±2.1; 13.6 (11.6–14.6) |
| Platelets<br>(×10<sup>9</sup>/L; normal range 150–400) | 210.8±0.02; 200 (162–247) | 207±69.2; 200 (162–240) | 214.9±90.3; 199.5 (160–264) |
| Alanine aminotransferase *<br>(U/L; normal range < 51) | 35.9±41.7; 26 (17–41) | 31.45±25.9; 24 (16–41) | 40.6±53.4; 26 (18–44) |
| Aspartate aminotransferase * | 43.2±36.5; 33 (25–49) | 37.6±21.9; 31 (24–44) | 49.1±46.8; 36.5 (26–61) |
| (U/L; normal range < 51) |  |  |  |
| Gamma-glutamyl transpeptidase<br>(U/L; normal range < 55) | 72.6±80.3; 42 (22–78) | 72.9±78.3; 46 (23–81) | 72.3±82.7; 36.5 (22–78) |
| Alkaline phosphatase<br>(U/L; normal range 40–150) | 97.96±47.1; 85 (65–117) | 97.7±42; 88 (65–117) | 98.2±52.2; 83.5 (65–118) |
| Total bilirubin<br>(mg/dL; normal range 0.3–1.2) | 0.7±0.4; 0.6 (0.5–0.9) | 0.69±0.36; 0.6 (0.4–0.8) | 0.8±0.5; 0.6 (0.5–0.9) |
| Creatine kinase<br>(U/L; normal range < 172) | 186.4±268.2; 97 (61–180) | 169.4±263; 94 (61–168) | 204.4±274.1; 111 (58–207) |
| Lactate dehydrogenase<br>(U/L; normal range < 248) | 310.6±115.5; 291 (240–370) | 267.2±81.98; 253 (199–305) | 356.5±127.97; 334.5 (286–414) |
| Serum creatinine<br>(mg/dL; normal range 0.67–1.17) | 1.3±1.03; 0.92 (0.8–1.22) | 1.34±1.23; 0.91 (0.8–1.17) | 1.17±0.79; 0.95 (0.71–1.24) |
| Troponin-I<br>(ng/L; normal range 1–35) | 133.7±1307.97; 10.4 (5.7–22.9) | 35±87.3; 9.5 (4.8–22.9) | 238.2±1874.2; 11.1 (6.6–24.3) |
| D-dimer<br>(µg/mL; normal range 0.2–0.35) | 1783±5486; 436 (267–868)<br>(n=176) | 2056.98±6699.7; 415 (254–868) | 1495.6±3848.9; 461.5 (291–845) |
| Fibrinogen *<br>(mg/dL; normal range 160–400) | 568.4±181.6; 576 (436–684) | 533.3±158.7; 542 (414–625) | 605.7±197.1; 608 (451–719) |
| Triglycerides<br>(mg/dL; normal range 10–150) | 114.8±45.7; 106 (82–139) | 114.2±50.6; 106 (78–136) | 115.4±40.1; 107 (83–139) |
| *Inflammation Biomarkers* |  |  |  |
| C-reactive protein ***<br>(mg/dL; normal range < 1.0) | 7.9±7.1; 6.3 (2.2–11.4)<br>(n=176) | 5.37±5.3; 3.98 (1.38–8.01)<br>(n=90) | 10.59±7.76; 10.2 (3.89–15.3) |
| C-reactive protein log ***<br>(per 1 mg dl<sup>-1</sup> increase)<sup>a</sup> | 0.7±0.5; 0.8 (0.34–1.06)<br>(n=176) | 0.47±0.55; 0.6 (0.14–0.9)<br>(n=90) | 0.85±0.45; 1.01 (0.59–1.19) |
| Interleukin-6<br>(pg/mL; normal range < 6.4) | 57.8±71.6; 38 (20–68) | 54.97±82.16; 30 (18–51) | 60.8±58.7; 49.5 (23–86) |
| IL-6 log<br>(per 1 pg ml<sup>-1</sup> increase)<sup>a</sup> | 1.5±0.43; 1.58 (1.3–1.8) | 1.5±0.42; 1.48 (1.25–1.71) | 1.62±0.4; 1.7 (1.36–1.93) |
| Pentraxin 3 ***<br>(ng/mL) | 37.4±44.96; 18.5 (8.9–48.4) | 27.1±34.95; 14.45 (6.85–32.1) | 48.2±51.6; 28.7 (14.9–67.7) |
| Pentraxin 3 log***<br>(per 1 ng ml<sup>-1</sup> increase)<sup>a</sup> | 1.3±0.5; 1.3 (0.95–1.68) | 1.16±0.5; 1.16 (0.84–1.51) | 1.489±0.44; 1.46 (1.17–1.83) |
| Procalcitonin *<br>(ng/mL; normal range 0.05–0.5) | 0.4±0.72; 0.14 (0.08–0.365)<br>(n=176) | 0.3±0.45; 0.12 (0.07–0.29)<br>(n=90) | 0.49±0.91; 0.19 (0.1–0.52) |
| Ferritin<br>(ng/mL; normal range 23.9–336.2) | 537.3±585.9; 347 (174.8–722) | 468.1±615.6; 309.9 (148–624.6) | 610.6±546.91; 481 (174.8–902.2) |

\* Statistical significance between MILD COVID-19 subgroups (QCT< and ≥ 9.5%): \* p<0.05; \*\* p<0.01; \*\*\* p<0.001.
<sup>a</sup> Variable expressed in logarithmic scale.

### Association with radiological lung damage

Investigating correlations between QCT measurement of lung damage and all the variables, some laboratory characteristics were associated with radiological deterioration on univariable analysis (Table 2). In particular, hemoglobin, platelets, AST, LDH, fibrinogen, CRP (log scale), and PTX3 (log scale) were significantly correlated with CT scan in hospitalized COVID-19 patients. On the contrary, all the other demographic features, comorbidities, severity-related biomarkers, laboratory characteristics, and inflammatory indexes were not associated with an increased risk. Of note, neither IL-6 nor Ferritin were significant. After the selection of variables, as described in materials and methods section, multivariable analysis demonstrated a correlation between PTX3, CRP and LDH and radiological lung damage (compromised lung ≥9·5%) (Table 2.)

**Table 2.**
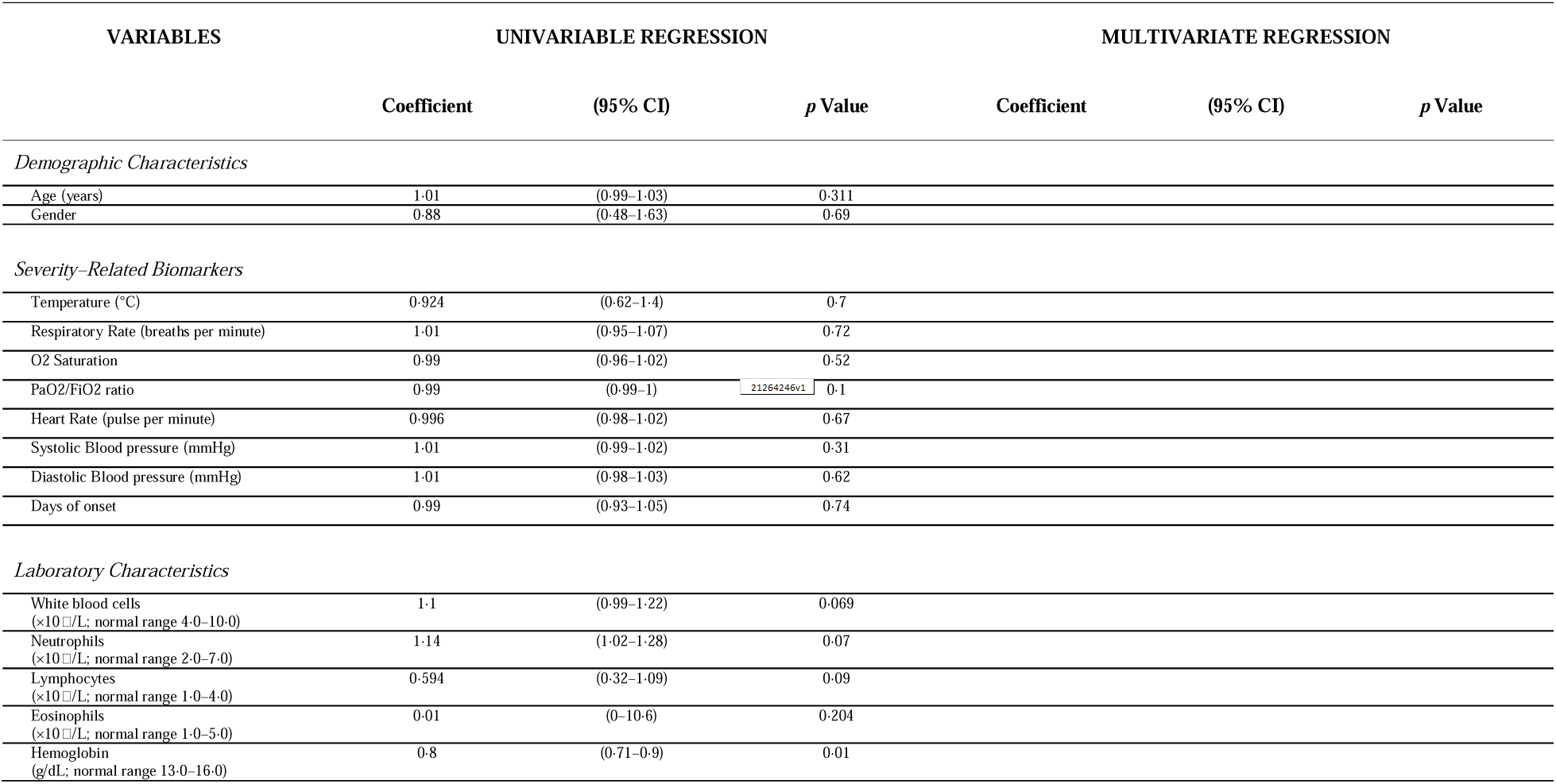
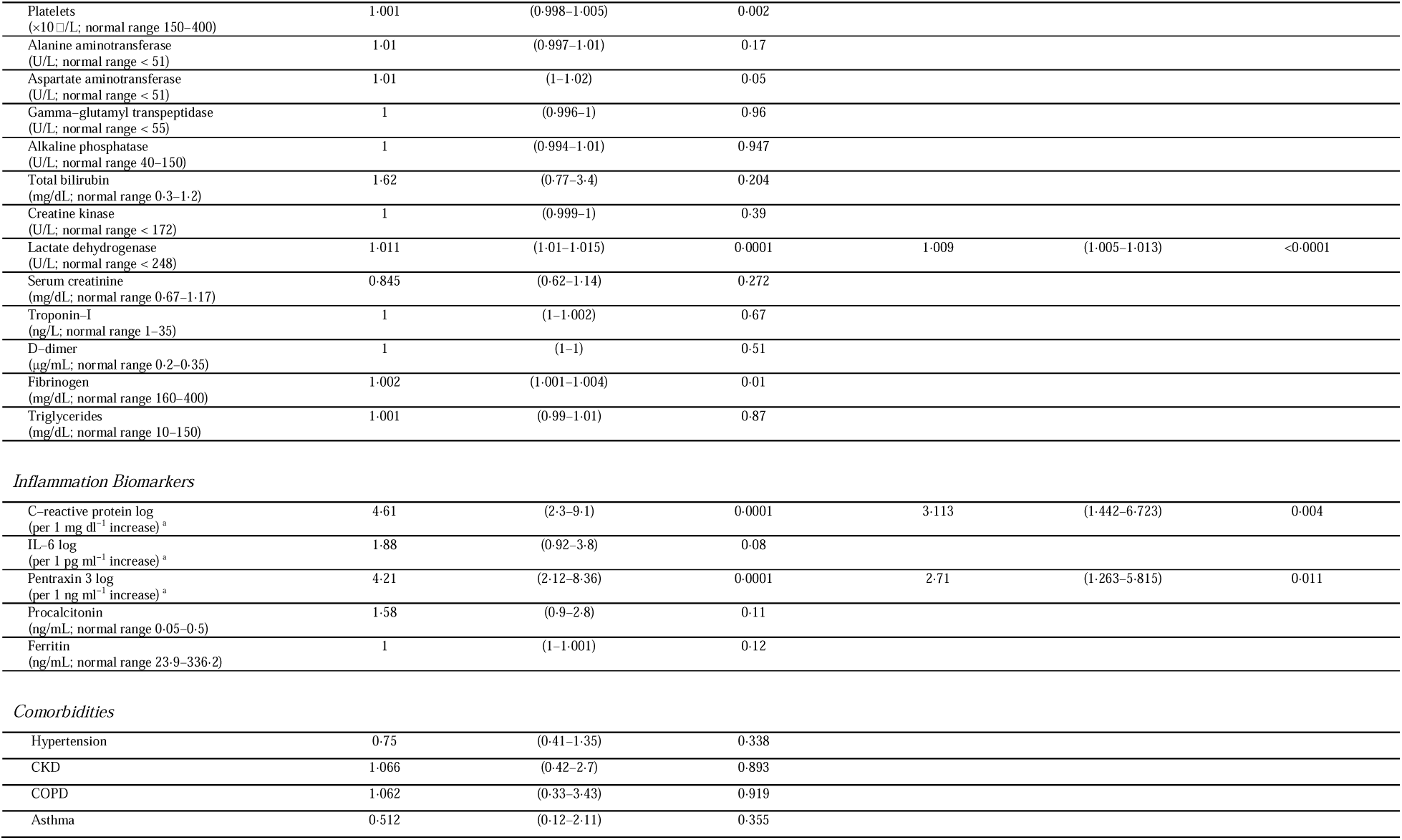
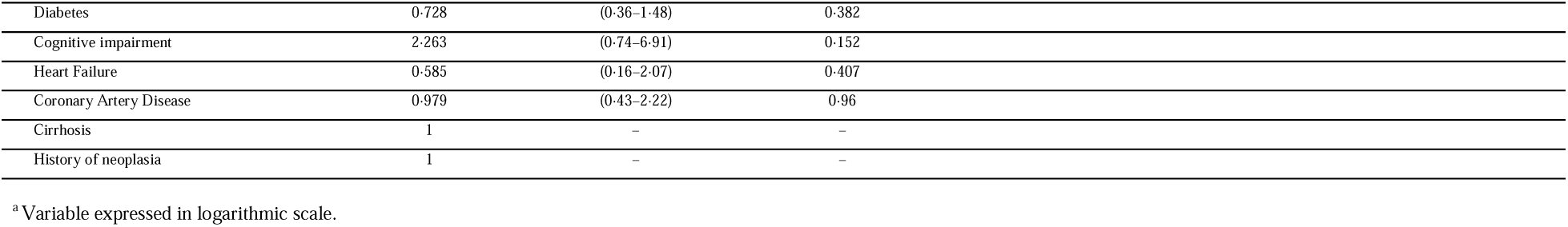
Univariate and Multivariate regressions in MILD COVID-19 (patients with PaO2/FiO2 ratio greater than 300).

| VARIABLES | UNIVARIABLE REGRESSION |  |  | MULTIVARIATE REGRESSION |  |  |
| --- | --- | --- | --- | --- | --- | --- |
|  | Coefficient | (95% CI) | *p* Value | Coefficient | (95% CI) | *p* Value |
| *Demographic Characteristics* |  |  |  |  |  |  |
| Age (years) | 1.01 | (0.99–1.03) | 0.311 |  |  |  |
| Gender | 0.88 | (0.48–1.63) | 0.69 |  |  |  |
| *Severity–Related Biomarkers* |  |  |  |  |  |  |
| Temperature (°C) | 0.924 | (0.62–1.4) | 0.7 |  |  |  |
| Respiratory Rate (breaths per minute) | 1.01 | (0.95–1.07) | 0.72 |  |  |  |
| O2 Saturation | 0.99 | (0.96–1.02) | 0.52 |  |  |  |
| PaO2/FiO2 ratio | 0.99 | (0.99–1) | 0.1 |  |  |  |
| Heart Rate (pulse per minute) | 0.996 | (0.98–1.02) | 0.67 |  |  |  |
| Systolic Blood pressure (mmHg) | 1.01 | (0.99–1.02) | 0.31 |  |  |  |
| Diastolic Blood pressure (mmHg) | 1.01 | (0.98–1.03) | 0.62 |  |  |  |
| Days of onset | 0.99 | (0.93–1.05) | 0.74 |  |  |  |
| *Laboratory Characteristics* |  |  |  |  |  |  |
| White blood cells<br>(×10<sup>9</sup>/L; normal range 4.0–10.0) | 1.1 | (0.99–1.22) | 0.069 |  |  |  |
| Neutrophils<br>(×10<sup>9</sup>/L; normal range 2.0–7.0) | 1.14 | (1.02–1.28) | 0.07 |  |  |  |
| Lymphocytes<br>(×10<sup>9</sup>/L; normal range 1.0–4.0) | 0.594 | (0.32–1.09) | 0.09 |  |  |  |
| Eosinophils<br>(×10<sup>9</sup>/L; normal range 1.0–5.0) | 0.01 | (0–10.6) | 0.204 |  |  |  |
| Hemoglobin<br>(g/dL; normal range 13.0–16.0) | 0.8 | (0.71–0.9) | 0.01 |  |  |  |
| Platelets<br>($\times 10^9$/L; normal range 150–400) | 1.001 | (0.998–1.005) | 0.002 | | | |
| Alanine aminotransferase<br>(U/L; normal range < 51) | 1.01 | (0.997–1.01) | 0.17 |  |  |  |
| Aspartate aminotransferase<br>(U/L; normal range < 51) | 1.01 | (1–1.02) | 0.05 |  |  |  |
| Gamma–glutamyl transpeptidase<br>(U/L; normal range < 55) | 1 | (0.996–1) | 0.96 |  |  |  |
| Alkaline phosphatase<br>(U/L; normal range 40–150) | 1 | (0.994–1.01) | 0.947 |  |  |  |
| Total bilirubin<br>(mg/dL; normal range 0.3–1.2) | 1.62 | (0.77–3.4) | 0.204 |  |  |  |
| Creatine kinase<br>(U/L; normal range < 172) | 1 | (0.999–1) | 0.39 |  |  |  |
| Lactate dehydrogenase<br>(U/L; normal range < 248) | 1.011 | (1.01–1.015) | 0.0001 | 1.009 | (1.005–1.013) | <0.0001 |
| Serum creatinine<br>(mg/dL; normal range 0.67–1.17) | 0.845 | (0.62–1.14) | 0.272 |  |  |  |
| Troponin-I<br>(ng/L; normal range 1–35) | 1 | (1–1.002) | 0.67 |  |  |  |
| D–dimer<br>(μg/mL; normal range 0.2–0.35) | 1 | (1–1) | 0.51 | | | |
| Fibrinogen<br>(mg/dL; normal range 160–400) | 1.002 | (1.001–1.004) | 0.01 |  |  |  |
| Triglycerides<br>(mg/dL; normal range 10–150) | 1.001 | (0.99–1.01) | 0.87 |  |  |  |
| *Inflammation Biomarkers* |  |  |  |  |  |  |
| C-reactive protein log<br>(per 1 mg dl<sup>-1</sup> increase)<sup>a</sup> | 4.61 | (2.3–9.1) | 0.0001 | 3.113 | (1.442–6.723) | 0.004 |
| IL-6 log<br>(per 1 pg ml<sup>-1</sup> increase)<sup>a</sup> | 1.88 | (0.92–3.8) | 0.08 |  |  |  |
| Pentraxin 3 log<br>(per 1 ng ml<sup>-1</sup> increase)<sup>a</sup> | 4.21 | (2.12–8.36) | 0.0001 | 2.71 | (1.263–5.815) | 0.011 |
| Procalcitonin<br>(ng/mL; normal range 0.05–0.5) | 1.58 | (0.9–2.8) | 0.11 |  |  |  |
| Ferritin<br>(ng/mL; normal range 23.9–336.2) | 1 | (1–1.001) | 0.12 |  |  |  |

*Comorbidities*
|  |  |  |  |
| --- | --- | --- | --- |
| Hypertension | 0.75 | (0.41–1.35) | 0.338 |
| CKD | 1.066 | (0.42–2.7) | 0.893 |
| COPD | 1.062 | (0.33–3.43) | 0.919 |
| Asthma | 0.512 | (0.12–2.11) | 0.355 |

|  |  |  |  |
| --- | --- | --- | --- |
| Diabetes | 0.728 | (0.36–1.48) | 0.382 |
| Cognitive impairment | 2.263 | (0.74–6.91) | 0.152 |
| Heart Failure | 0.585 | (0.16–2.07) | 0.407 |
| Coronary Artery Disease | 0.979 | (0.43–2.22) | 0.96 |
| Cirrhosis | 1 | – | – |
| History of neoplasia | 1 | – | – |

<sup>a</sup> Variable expressed in logarithmic scale.

Based on these results, goodness of fit of the model was then measured with cvAUC obtaining a level of accuracy of 0·794±0·107 (CI 95%: 0·74–0·87) (Figure 1). Other models, tested with different combinations of variables, resulted less accurate than the chosen version (Table 3). The other patient subgroups were not tested because of the radiological lung injury results greater than the established outcome represented by the 9·5% threshold.

**Figure 1.**
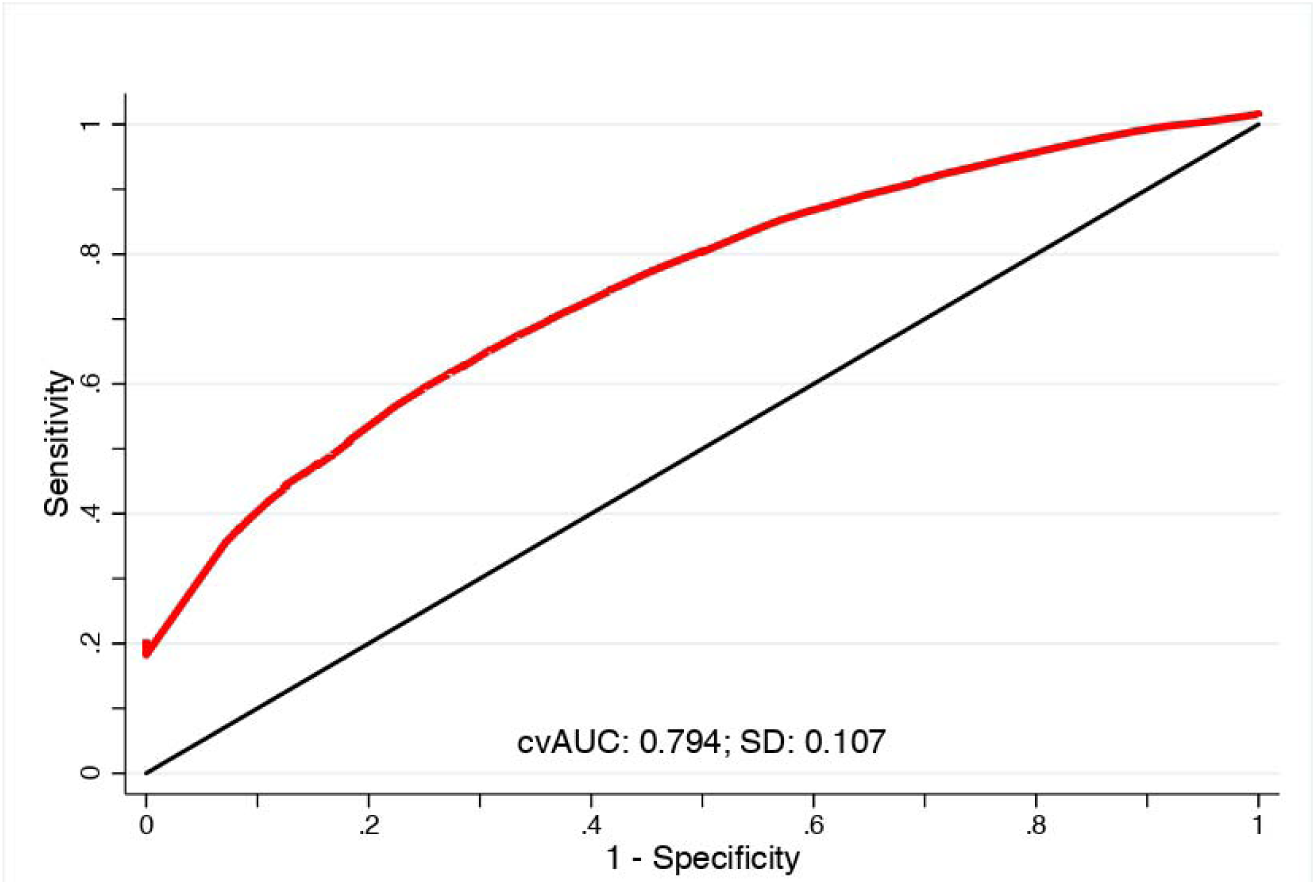
Predictive model Cross-validated Area Under the ROC Curve (cvAUC) performed on the MILD COVID-19 group of patients. The result demonstrates a level of accuracy of 0·794±0·107 (CI 95%: 0·74–0·87) in predicting radiological injury measured by QCT and represented in binary form (the selected threshold of 9·5% represents the amount of lung parenchyma involved by the SARS-CoV2 pneumonia which is predictive of clinical deterioration).

**Table 3.** Model accuracy and sensitivity analysis. The selected model demonstrates the best accuracy level; indeed, other combinations of the three variables don’t reach the same performance suggesting specific biological implication in the SARS-CoV2 infections by the three different biomarkers.

| Variables | Coefficient | (95% CI) | *p* Value | Accuracy | (95% CI) |
| --- | --- | --- | --- | --- | --- |
| Pentraxin 3 log<br>(per 1 ng ml<sup>-1</sup> increase)<sup>a</sup> | 2.71 | (1.263–5.815) | 0.011 |  |  |
| C-reactive protein log<br>(per 1 mg dl<sup>-1</sup> increase)<sup>a</sup> | 3.113 | (1.442–6.723) | 0.004 | 0.794±0.107 | 0.74–0.87 |
| Lactate dehydrogenase<br>(U/L; normal range < 248) | 1.009 | (1.005–1.013) | 0.0001 |  |  |
| C-reactive protein log<br>(per 1 mg dl<sup>-1</sup> increase)<sup>a</sup> | 3.62 | (1.72–7.61) | 0.001 | 0.773±0.107 | 0.705–0.846 |
| Lactate dehydrogenase<br>(U/L; normal range < 248) | 1.01 | (1.005–1.01) | 0.0001 |  |  |
| Pentraxin 3 log<br>(per 1 ng ml<sup>-1</sup> increase)<sup>a</sup> | 3.409 | (1.62–7.18) | 0.001 | 0.7799±0.086 | 0.697–0.843 |
| Lactate dehydrogenase<br>(U/L; normal range < 248) | 1.01 | (1.01–1.01) | 0.0001 |  |  |
| Pentraxin 3 log<br>(per 1 ng ml<sup>-1</sup> increase)<sup>a</sup> | 3.02 | (1.49–6.13) | 0.002 | 0.725±0.106 | 0.673–0.822 |
| C-reactive protein log<br>(per 1 mg dl<sup>-1</sup> increase)<sup>a</sup> | 3.754 | (1.84–7.66) | 0.0001 |  |  |

<sup>a</sup> Variable expressed in logarithmic scale.

### Score

A preliminary tool to predict significant radiological involvement has been developed. We provided easy-to-apply instructions to derive the score for each patient given his/her PTX3, CRP and LDH levels (Table 4). The prognostic index showed high predictive accuracy as demonstrated by cvAUC of 0·801±0·084 (CI 95%, 0·75–0·88) (Figure 2, Panel A). The score performance was then assessed on the total subgroup of paucisymptomatic COVID-19, including septic and neoplastic patients previously excluded (226 pts) obtaining cvAUC of 0·762±0·093 (CI 95%, 0·65–0·797) (Figure 2, Panel B).

**Table 4.** CT-Lung involvement prediction index scoring algorithm.

| Category | Index cut-offs | Risk level |
| --- | --- | --- |
| Lung-CT involvement > 9.5% | 1 | 5% |
|  | 3 | 30% |
|  | 5 | 80% |
|  | 8 | 99% |

**Figure 2.**
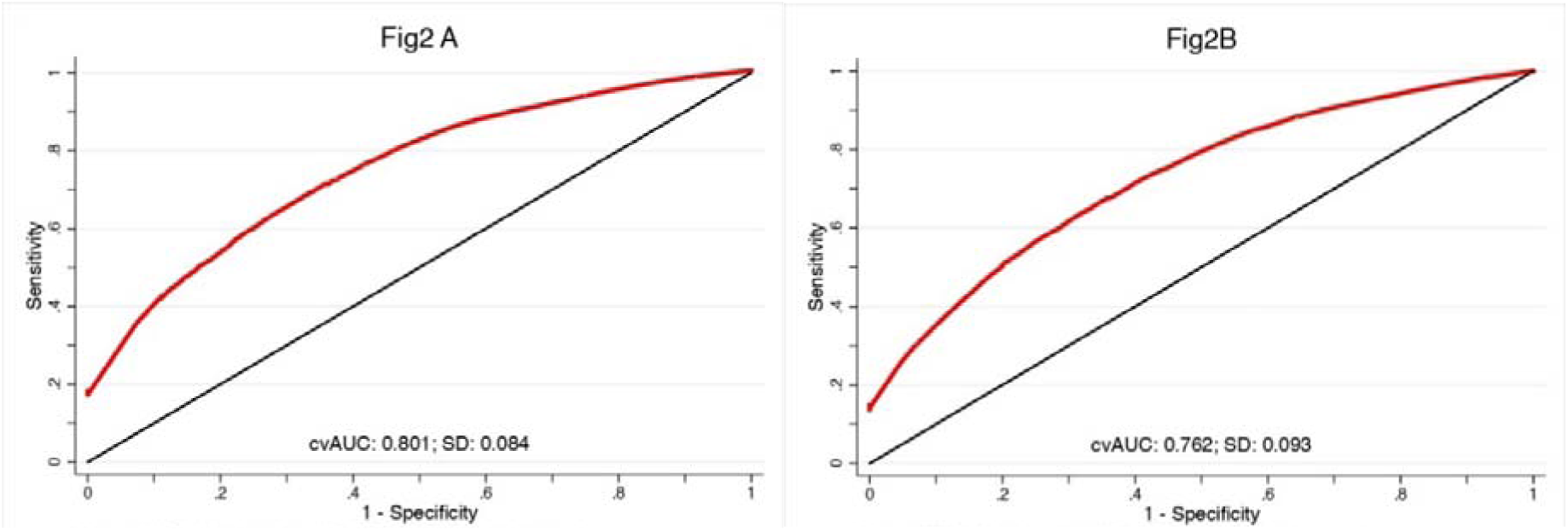
Panel A: Score Cross-validated Area Under the ROC Curve (cvAUC). The figure shows the score performance on the MILD COVID-19 group (177 patients) with a result of a 0·801±0·084 accuracy (CI 95%, 0·75– 0·88). Panel B: Total cohort predictive model Cross-validated Area Under the ROC Curve (cvAUC). Score utility in real life was assessed on the totality of paucisymptomatic COVID-19, considering also septic and neoplastic patients previously excluded (226 patients) obtaining cvAUC of 0·762±0·093 (CI 95%, 0·65–0·797).

### Clinical worsening

Clinical deterioration was identified as a progression to ICU ward and the occurrence of death. In the group of patients with PaO2/FiO2 ratio ≥300, 14 patients were transferred to intensive care regimen and among those, 2 died. Instead, the overall deaths in this group counted 21 patients (11·9%). We analysed the correlation between the radiological compromised lung and a composite outcome characterized by ICU transfer or death. Both the PaO2/FiO2≥300 group (OR 3·95; p<0·001, CI 95%, 1·73–9·04) and PaO2/FiO2<300 (OR 7·03; p<0·011; CI 95%, 1·57–31·4) resulted significant. The correlation between our multivariate model (including PTX3, CRP and LDH) and clinical outcome was analysed demonstrating the association of PTX3 and LDH in the PaO2/FiO2≥300 subgroup (Table 5). For the most compromised category (PaO2/FiO2<300) the analysis identified LDH as significantly associated with ICU/death (Table 5).

**Table 5.** Progression to ICU/death. The clinical composite outcome (ICU/death) was correlated with our model (including PTX3, CRP and LDH) demonstrating a significant association of PTX3 and LDH in the MILD COVID-19 group; instead in SEVERE COVID-19 only LDH was relevant.

| Patient category | Number of pts | Variables | OR | (95% CI) | *p* Value |
| --- | --- | --- | --- | --- | --- |
| MILD COVID-19<br>PaO<sub>2</sub>/FiO<sub>2</sub>≥300 | 177 | Pentraxin 3 log<br>(per 1 ng ml<sup>-1</sup> increase)<sup>a</sup> | 3.46 | (1.367-8.779) | 0.009 |
|  |  | C-reactive protein log<br>(per 1 mg dl<sup>-1</sup> increase)<sup>a</sup> | 0.87 | (0.37-2.07) | 0.761 |
|  |  | Lactate dehydrogenase<br>(U/L; normal range < 248) | 1.005 | (1.001-1.01) | 0.007 |
| SEVERE COVID-19<br>PaO<sub>2</sub>/FiO<sub>2</sub><300 | 189 | Pentraxin 3 log<br>(per 1 ng ml<sup>-1</sup> increase)<sup>a</sup> | 0.575 | (0.3-1.1) | 0.096 |
|  |  | C-reactive protein log<br>(per 1 mg dl<sup>-1</sup> increase)<sup>a</sup> | 2.68 | (0.85-8.42) | 0.092 |
|  |  | Lactate dehydrogenase<br>(U/L; normal range < 248) | 1.004 | (1.002-1.006) | 0.0001 |

<sup>a</sup> Variable expressed in logarithmic scale.

## Discussion

The results presented here show that LDH, CRP and PTX3 together correlate with QCT lung damage in paucisymptomatic COVID-19 patients. In particular, in early disease progression dosing the markers provides a footprint of radiological findings with a high level of accuracy (0·794±0·107; CI 95%: 0·74–0·87). Since the analysis refers to a specific QCT threshold significant for oxygen need and hospitalization, the score reflects the probability of clinical worsening in mild COVID-19. Moreover, the data show an unexpected 11·9% of mortality in paucisymptomatic patients highlighting clinical needs in early assessment of this condition. The proposed tool was analyzed demonstrating a significant correlation with clinical deterioration in every subgroup of patients.

The present study has limitations. It is retrospective and single-centre and needs to be extended on a large scale to be validated. Moreover, the significant association was defined in a selected population of paucisymptomatic patients at admission, but we missed the follow up during the hospitalization because of the lack of standardized time in lung CT. It will therefore be important to confirm the value with external cohorts of patients considering time and data for the radiologic follow-up.

The rapid dissemination of SARS-CoV-2 globally and the outbreak of new viral variants compels the evolution of new tools to approach evolving clinical and social needs. Even though the start of vaccination, the spread of the virus remains high as demonstrated by the growing pandemic which counts more than 200 million of people infected worldwide with 4 million of deaths (WHO report 6 August 2021). With these assumptions, it becomes crucial to develop innovative instruments to manage every clinical condition the virus determines with the aim to be applied on a large scale. Nowadays, a major part of the studies on COVID-19 are focused on a better definition of severe and critically ill patients to define new strategies of clustering and discover additional biomarkers. The most diffuse approach is based on complex multiparametric scores which rely on clinical and laboratory variables leading to esteem deterioration of health status and mortality.^39,40^ Other methods focus on the identification of specific serologic signatures representing organ damage or definite clinical complications. Some of these put the emphasis on cardiac injury^41,42^ as well as on thromboembolic phenomena,^43,44^ since they are considered some of the most threatening factors which precipitate the health status. Recently, endothelial cells infection and metabolic disruption have been considered some of the most harmful pathological events due to the failure of alveolar-capillary barrier. Vascular inflammation, dysfunction and damage were assessed in several studies by determining serological biomarkers which demonstrate distinct relation to clinical worsening and mortality.^9,45^ Nevertheless constant attempts to develop new measures of severity and to assist clinicians, to date there are no options predicting deterioration in mild COVID-19 patients as well as response to therapies. Paucisymptomatic subjects represent a challenge for health systems because of the presence of minimal alterations at vital signs, laboratory tests and blood gases analysis which lead to underestimate the clinical picture. Considering COVID-19 as an evolving condition, it became clear how first assessment in an ER setting could overlook patients who are prone to deteriorate, especially at the beginning of the disease. Since the absence of biomarkers, clinicians experience uncertainty during the process to identify subjects needed to be monitored or promptly treated; indeed, as demonstrated by our data, almost 12% of mild COVID-19 died underlining how variable this condition could be. For these reasons, the presented model represents an attempt to improve first assessment of people tested positive with normal vital signs offering a new instrument to predict unfavourable evolution. Moreover, this could impact positively in triaging patients leading to evaluate the extension of pneumonia without undergoing to radiological procedures.

The reason(s) for the strong correlation between combined measurement of PTX3, CRP and LDH and QCT disease assessment and outcome remains a matter of speculation. CRP is produced in the liver mainly in response to IL-6 and therefore reflects a systemic amplification loop of inflammation. The major sources of PTX3 in COVID-19 are myeloid cells and endothelial cells^18^ and therefore may serve as direct marker of involvement of prime cellular drivers of inflammation at the tissue level. LDH is a marker of cell and tissue damage. Therefore CRP, PTX3 and LDH may represent complementary biomarkers reflecting local and systemic inflammation and tissue damage.

In conclusion, there is a growing need to cluster COVID-19 patients identifying those prone to clinical worsening in spite of mild symptoms and normal functional tests (id PaO2/FiO2). This clinical necessity could at least in part be addressed by the serological markers analysed in the present study which represent a fingerprint of SARS-CoV-2 pathological mechanisms. The proposed predictive model considers pneumocytes lysis, but also summarizes the different extent of local and systemic inflammatory cascade reflecting to a considerable extent cardiologic damage.

## Supporting information

Letter

Humanitas COVID-19 Task Force

## Data Availability

The clinical data that support the findings of this study are available in Tables 1-6, Figure s1 and from the corresponding authors upon request.

## Acknowledgement

This work was supported by a philanthropic donation by Dolce & Gabbana fashion house (to A.M., C.G.) and by a grant from Italian Ministry of Health for COVID-19 (to A.M. and C.G.).

This work was conducted in the framework of, and made possible by, the collective effort of the Humanitas COVID-19 Task Force (the members are listed in the Supplementary Note).

## SUPPLEMENTARY MATERIALS

**Figure S1.**
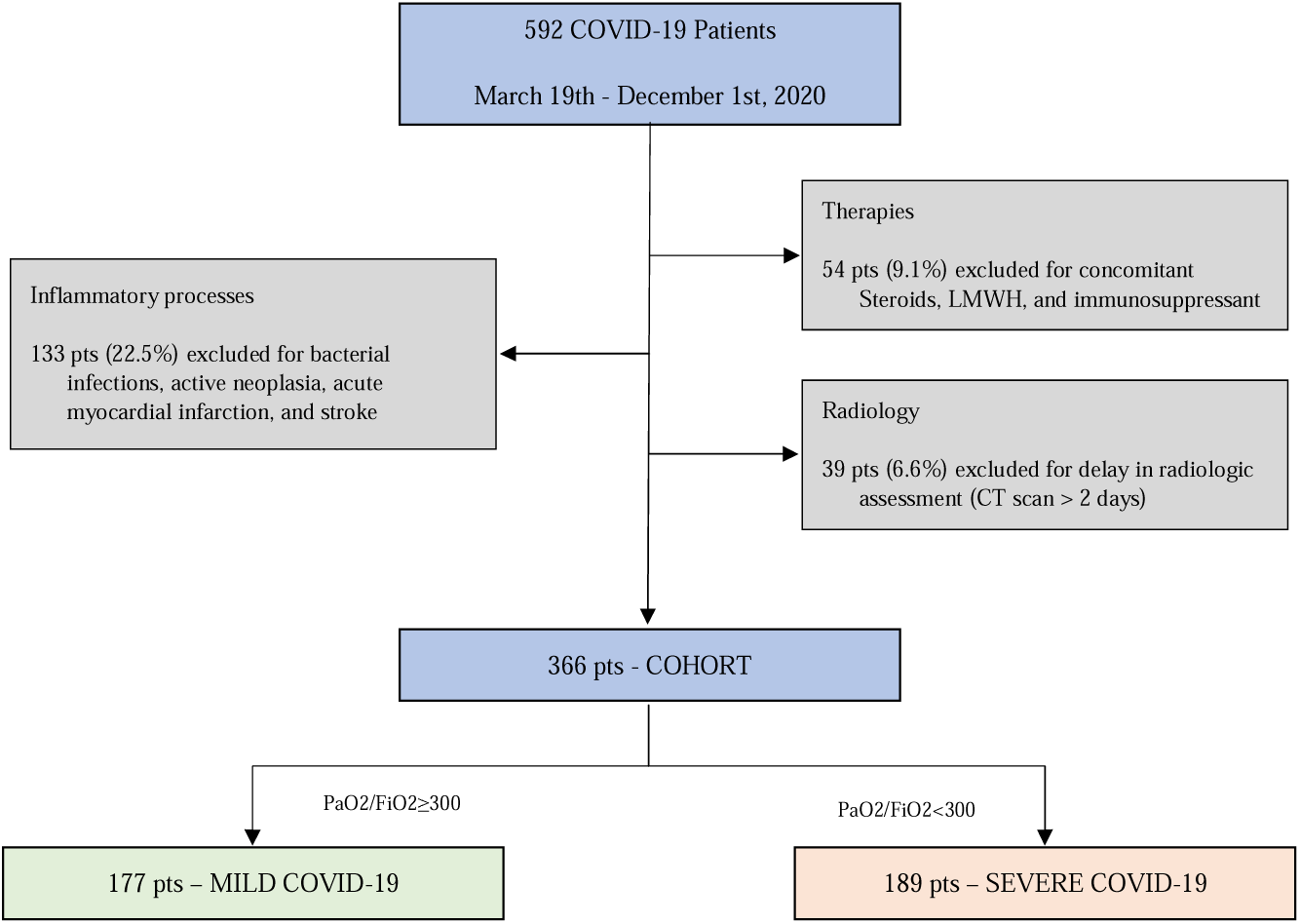
Flow Chart of the enrolled patients. 592 subjects diagnosed with COVID-19 and admitted to Humanitas Research Hospital between March 19th and December 1st 2020 were considered. Confounders for the study (grey squares) were identified as follow: 1) inflammatory processes which can influence the levels of dosed inflammation biomarkers (bacterial infections, active neoplasia, and acute pathological conditions such as acute myocardial infarction and stroke); 2) therapies that can interfere on natural history of disease as well as immune system responsiveness (i.e. steroids, LMWH and immunosuppressant); 3) radiology, CT scan performed after 2 days were excluded (39, 6·6%) since radiological features were not considered representative of the clinical and blood tests assessment. The resulting cohort is represented by 366 patients, clustered based on severity in MILD COVID-19 (paucisymptomatic patients with PaO2/FiO2≥300, n.177; green square) and SEVERE COVID-19 (symptomatic patients with PaO2/FiO2<300, n.189, red square).

**Table S1.**
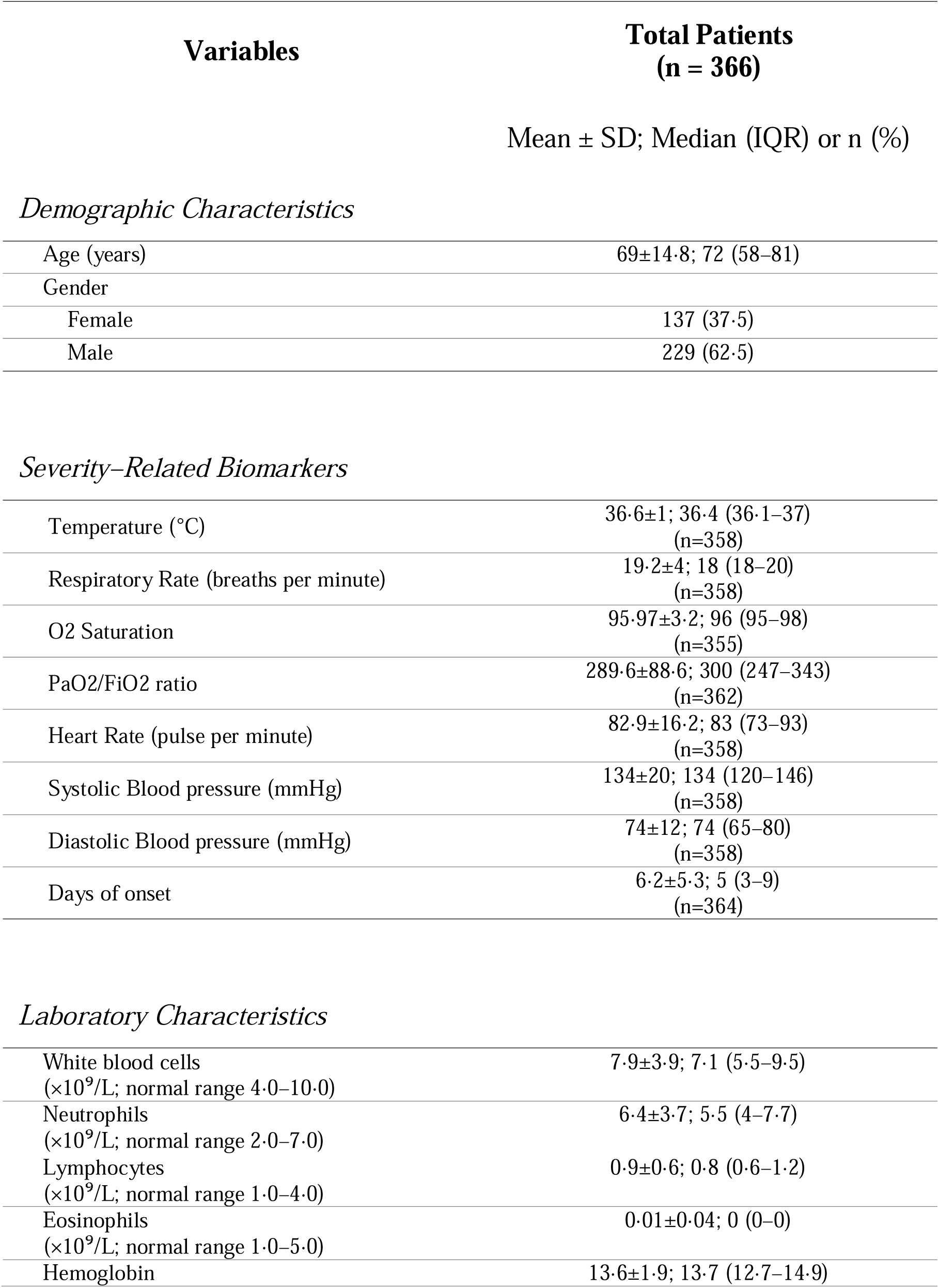
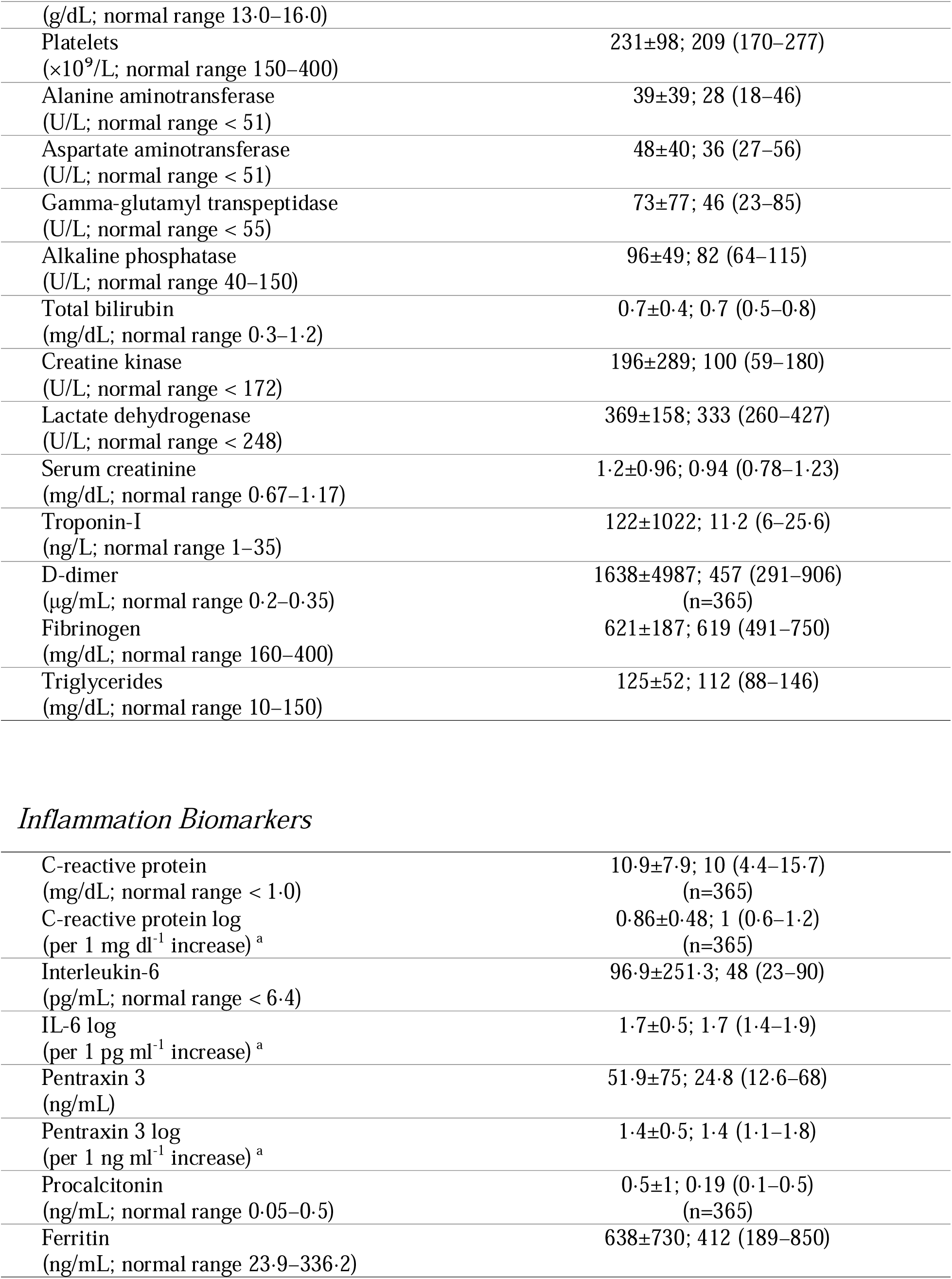
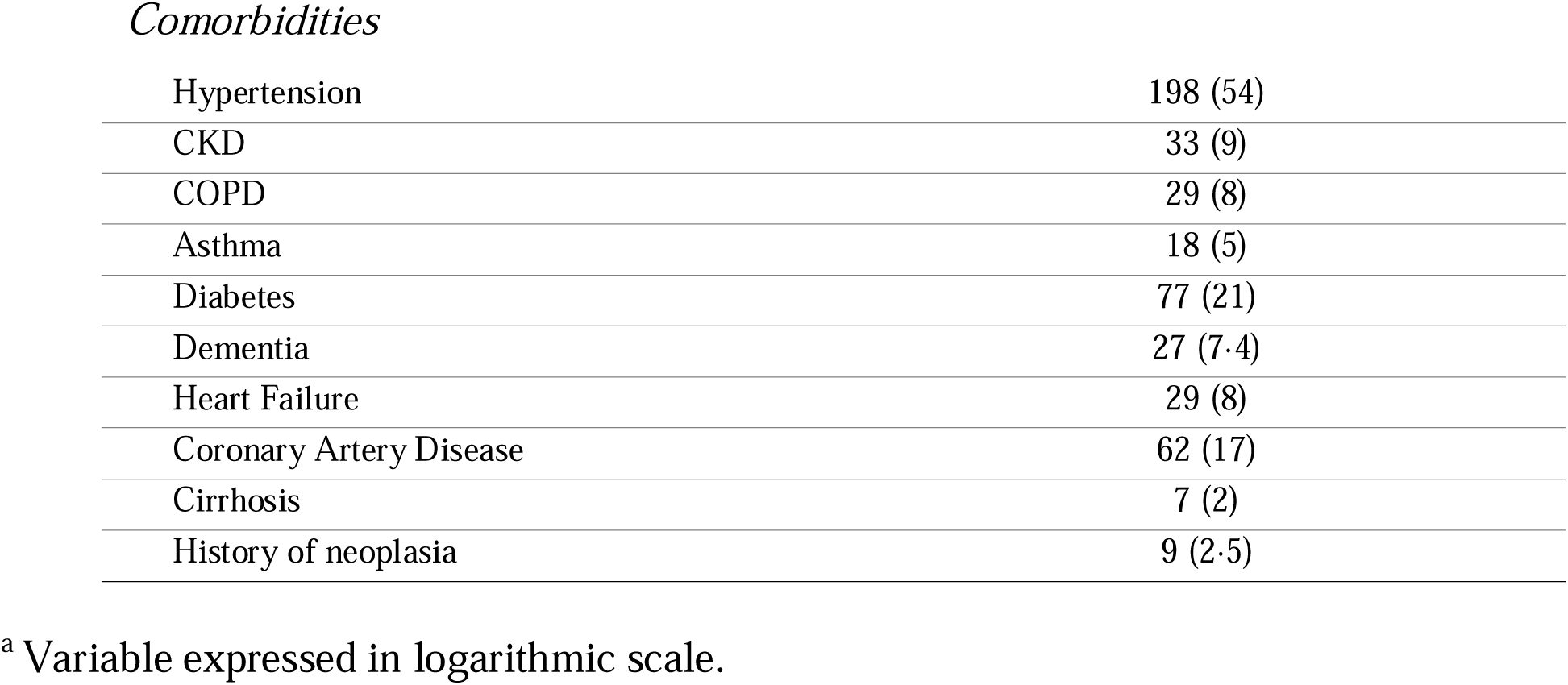
Demographics, severity-related and blood test of the total cohort.

| Variables | Total Patients<br>(n = 366) |
| --- | --- |
|  | Mean ± SD; Median (IQR) or n (%) |
| *Demographic Characteristics* |  |
| Age (years) | 69±14.8; 72 (58–81) |
| Gender |  |
| Female | 137 (37.5) |
| Male | 229 (62.5) |
| *Severity-Related Biomarkers* |  |
| Temperature (°C) | 36.6±1; 36.4 (36.1–37)<br>(n=358) |
| Respiratory Rate (breaths per minute) | 19.2±4; 18 (18–20)<br>(n=358) |
| O2 Saturation | 95.97±3.2; 96 (95–98)<br>(n=355) |
| PaO2/FiO2 ratio | 289.6±88.6; 300 (247–343)<br>(n=362) |
| Heart Rate (pulse per minute) | 82.9±16.2; 83 (73–93)<br>(n=358) |
| Systolic Blood pressure (mmHg) | 134±20; 134 (120–146)<br>(n=358) |
| Diastolic Blood pressure (mmHg) | 74±12; 74 (65–80)<br>(n=358) |
| Days of onset | 6.2±5.3; 5 (3–9)<br>(n=364) |
| *Laboratory Characteristics* |  |
| White blood cells<br>(×10<sup>9</sup>/L; normal range 4.0–10.0) | 7.9±3.9; 7.1 (5.5–9.5) |
| Neutrophils<br>(×10<sup>9</sup>/L; normal range 2.0–7.0) | 6.4±3.7; 5.5 (4–7.7) |
| Lymphocytes<br>(×10<sup>9</sup>/L; normal range 1.0–4.0) | 0.9±0.6; 0.8 (0.6–1.2) |
| Eosinophils<br>(×10<sup>9</sup>/L; normal range 1.0–5.0) | 0.01±0.04; 0 (0–0) |
| Hemoglobin | 13.6±1.9; 13.7 (12.7–14.9) |
| (g/dL; normal range 13.0–16.0) |  |
| Platelets<br>($\times 10^9$/L; normal range 150–400) | 231±98; 209 (170–277) |
| Alanine aminotransferase<br>(U/L; normal range < 51) | 39±39; 28 (18–46) |
| Aspartate aminotransferase<br>(U/L; normal range < 51) | 48±40; 36 (27–56) |
| Gamma-glutamyl transpeptidase<br>(U/L; normal range < 55) | 73±77; 46 (23–85) |
| Alkaline phosphatase<br>(U/L; normal range 40–150) | 96±49; 82 (64–115) |
| Total bilirubin<br>(mg/dL; normal range 0.3–1.2) | 0.7±0.4; 0.7 (0.5–0.8) |
| Creatine kinase<br>(U/L; normal range < 172) | 196±289; 100 (59–180) |
| Lactate dehydrogenase<br>(U/L; normal range < 248) | 369±158; 333 (260–427) |
| Serum creatinine<br>(mg/dL; normal range 0.67–1.17) | 1.2±0.96; 0.94 (0.78–1.23) |
| Troponin-I<br>(ng/L; normal range 1–35) | 122±1022; 11.2 (6–25.6) |
| D-dimer<br>(μg/mL; normal range 0.2–0.35) | 1638±4987; 457 (291–906)<br>(n=365) |
| Fibrinogen<br>(mg/dL; normal range 160–400) | 621±187; 619 (491–750) |
| Triglycerides<br>(mg/dL; normal range 10–150) | 125±52; 112 (88–146) |
| *Inflammation Biomarkers* |  |
| C-reactive protein<br>(mg/dL; normal range < 1.0) | 10.9±7.9; 10 (4.4–15.7)<br>(n=365) |
| C-reactive protein log<br>(per 1 mg dl<sup>-1</sup> increase)<sup>a</sup> | 0.86±0.48; 1 (0.6–1.2)<br>(n=365) |
| Interleukin-6<br>(pg/mL; normal range < 6.4) | 96.9±251.3; 48 (23–90) |
| IL-6 log<br>(per 1 pg ml<sup>-1</sup> increase)<sup>a</sup> | 1.7±0.5; 1.7 (1.4–1.9) |
| Pentraxin 3<br>(ng/mL) | 51.9±75; 24.8 (12.6–68) |
| Pentraxin 3 log<br>(per 1 ng ml<sup>-1</sup> increase)<sup>a</sup> | 1.4±0.5; 1.4 (1.1–1.8) |
| Procalcitonin<br>(ng/mL; normal range 0.05–0.5) | 0.5±1; 0.19 (0.1–0.5)<br>(n=365) |
| Ferritin<br>(ng/mL; normal range 23.9–336.2) | 638±730; 412 (189–850) |
| *Comorbidities* |  |
| Hypertension | 198 (54) |
| CKD | 33 (9) |
| COPD | 29 (8) |
| Asthma | 18 (5) |
| Diabetes | 77 (21) |
| Dementia | 27 (7.4) |
| Heart Failure | 29 (8) |
| Coronary Artery Disease | 62 (17) |
| Cirrhosis | 7 (2) |
| History of neoplasia | 9 (2.5) |

<sup>a</sup> Variable expressed in logarithmic scale.

