## Supplementary material for "A PTX3/LDH/CRP signature correlates with lung injury CTs scan severity and disease progression in paucisymptomatic COVID-19": Letter

24 September 2021

Dear Editors,

Please find enclosed the manuscript entitled “*A PTX3/LDH/CRP signature correlates with lung injury CTs scan severity and disease progression in paucisymptomatic COVID-19*” to be submitted for publication to MedRxiv.

This paper reports that a serological signature of inflammation (PTX3, LDH and CRP) can serve as a low cost, low tech correlate of CT scan-assessed lung damage in COVID-19 paucisymptomatic patients.

The report is based on our previous discovery the PTX3 is a strong independent predictor of death in hospitalized patients, an observation confirmed in independent studies (e.g. ref. 18-21).

We speculate that these biomarkers may reflect different complementary components of the inflammatory response (systemic response, CRP; local myeloid and endothelial cell activation, PTX3; tissue damage, LDH) and that complementarity underlies their significance in relation to CT-scan assessed lung involvement.

Assessing disease severity and prognosis early in the natural history of COVID-19 represents a medical need and a challenge. The low tech low cost signature reported here may contribute to addressing this challenge and more so in deprived social contexts. Because of these considerations we feel that our report may deserve the attention of a prime journal.

We look forward to a critical assessment of our work.

Sincerely yours,

**Maurizio Cecconi, M.D.**

Head of Department Anaesthesia and Intensive Care
IRCCS Istituto Clinico Humanitas
Professor of Anaesthesia and Intensive Care
Humanitas University

**Alberto Mantovani, M.D.**

Scientific Director, Istituto Clinico Humanitas

Emeritus Professor, Humanitas University
