## Supplementary material for "A PTX3/LDH/CRP signature correlates with lung injury CTs scan severity and disease progression in paucisymptomatic COVID-19": Humanitas COVID-19 Task Force

| | |
|---|---|
| ANFRAY | CLEMENT |
| BELGIOVINE | CRISTINA |
| BERTOCCHI | ALICE |
| BOMBACE | SARA |
| BRESCIA | PAOLA |
| CALCATERRA | FRANCESCA |
| CALVI | MICHELA |
| CANCELLARA | ASSUNTA |
| CAPUCETTI | ARIANNA |
| CARENZA | CLAUDIA |
| CARLONI | SARA |
| CARNEVALE | SILVIA |
| CAZZETTA | VALENTINA |
| COIANIZ | NICOLÒ |
| DARWICH | ABBASS |
| DE PAOLI | FEDERICA |
| DI DONATO | RACHELE |
| DIGIFICO | ELISABETH |
| DURANTE | BARBARA |
| FARINA | FLORIANA MARIA |
| FERRARI | VALENTINA |
| FORNASA | GIULIA |
| FRANZESE | SARA |
| GIL GOMEZ | ANTONIO |
| GIUGLIANO | SILVIA |
| GOMES | ANA RITA |
| LIZIER | MICHELA |
| LO CASCIO | ANTONINO |
| MELACARNE | ALESSIA |
| MOZZARELLI | ALESSANDRO |
| MY | ILARIA |
| ORESTA | BIANCA |
| PASQUALINI | FABIO |
| PASTÒ | ANNA |
| PELAMATTI | ERICA |
| PERUCCHINI | CHIARA |
| POZZI | CHIARA |
| RIMOLDI | VALERIA |
| RIMOLDI | MONICA |
| SCARPA | ALICE |
| SILVESTRI | ALESSANDRA |
| SIRONI | MARINA |
| SPADONI | ILARIA |
| SPANO' | SALVATORE |
| SPATA | GIANMARCO |
| SUPINO | DOMENICO |

| | |
|---|---|
| TENTORIO | PAOLO |
| UMMARINO | ALDO |
| VALENTINO | SONIA |
| ZAGHI | ELISA |
| ZANON | VERONICA |
